# RaTEScore: A Metric for Radiology Report Generation

**DOI:** 10.1101/2024.06.24.24309405

**Authors:** Weike Zhao, Chaoyi Wu, Xiaoman Zhang, Ya Zhang, Yanfeng Wang, Weidi Xie

## Abstract

This paper introduces a novel, entity-aware metric, termed as **Ra**diological Report (**T**ext) **E**valuation (**RaTEScore**), to assess the quality of medical reports generated by AI models. RaTEScore emphasizes crucial medical entities, such as diagnostic outcomes and anatomical details. Moreover, it is robust against medical synonyms and sensitive to negation expressions. Technically, we developed a comprehensive medical NER dataset, **RaTE-NER**, and trained an NER model specifically for this purpose. This model enables the decomposition of complex radiological reports into constituent medical entities. The metric itself is derived by comparing the similarity of entity embeddings, obtained from a language model, based on their types and relevance to clinical significance. Our evaluations demonstrate that RaTEScore aligns more closely with human preference than existing metrics, validated both on established public benchmarks and our newly proposed **RaTE-Eval** benchmark.

## 1 Introduction

With the general advancement in natural language processing (OpenAI, 2023; Anil et al., 2023; Qiu et al., 2024; Wu et al., 2024) and computer vision (Li et al., 2023; Alayrac et al., 2022; OpenAI; Zhang et al., 2023; Wu et al., 2023a; Zhou et al., 2024), the pursuit of generalist medical artificial intelligence has grown increasingly promising and appealing (Moor et al., 2023; Wu et al., 2023b; Tu et al., 2024; Zheng et al., 2023; Zhao et al., 2023), leading to the development of free-text generative foundation models capable of understanding and interpreting radiology imaging studies. Yet, the complexity and specialized nature of clinical free-form texts, such as radiology reports and discharge summaries, present substantial challenges in evaluating model performance. There is an urgent need for a robust and lightweight free-text evaluation metric to better monitor the development of medical generative foundation models and drive advancements in the field of generalist medical artificial intelligence.

In the existing literature, four main types of metrics have been adopted to assess the similarity between free-form texts in medical scenarios, as shown in Figure 1. These include: (i) Word Overlap Metrics, such as BLEU (Papineni et al., 2002) and ROUGE (Lin, 2004). While intuitive, these metrics fail to capture negation or synonyms in sentences, thus neglecting the semantic factuality; (ii) Embedding Similarity Metrics, like BERTScore (Zhang et al., 2020), provide better semantic awareness but fail to emphasize key medical terms, leading to overlooking errors in critical conclusions; (iii) Metrics based on Named Entity Recognition (NER), such as RadGraph F1 (Yu et al., 2023a) and MEDCON (Yim et al., 2023). Although tailored for the medical domain, they struggle with synonym unification and are typically restricted to analyzing Chest X-ray reports; (iv) Metrics relying on large language models (LLMs) (Wei et al., 2024; Liu et al., 2023). While these metrics are better aligned with human preferences, they suffer from potential subjective biases and are prohibitively costly for large-scale evaluation.

**Figure 1:**
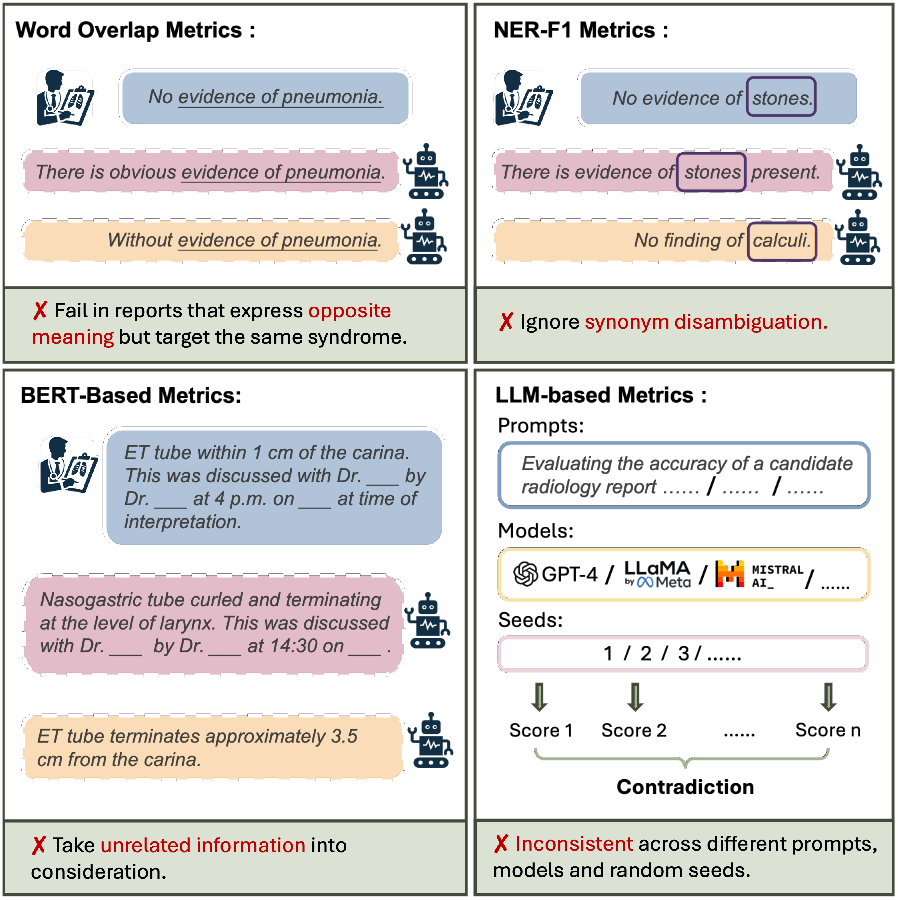
Existing evaluation metrics. We illustrate the limitations of current metrics. Blue boxes represent ground-truth reports; red and yellow boxes indicate correct and incorrect generated reports, respectively. The examples show that these metrics fail to identify opposite meanings and synonyms in the reports and are often disturbed by unrelated information.

In this study, we aim to develop a metric that prioritizes key medical entities such as diagnostic outcomes and anatomical features, while exhibiting robustness against complex medical synonyms and sensitivity to negation expressions. We present two contributions: First, we introduce **RaTEScore**, a novel evaluation metric specifically designed for radiology reports. This metric emphasizes entity-level assessments across various imaging modalities and body regions. Specifically, it starts by identifying medical entities and their types (*e*.*g*., anatomy, disease) from each sentence. To address the challenges associated with medical synonyms, we compute entity embeddings using a synonym disambiguation module and assess their cosine similarities. RaTEScore then calculates a final score based on weighted similarities that emphasize the clinical importance of the entity types involved.

Second, we have developed a comprehensive medical named-entity recognition (NER) dataset, **RaTE-NER**, which encompasses data from 9 modalities and 22 anatomical regions, derived from MIMIC-IV and Radiopaedia. In addition, we introduce **RaTE-Eval**, a novel benchmark designed to assess metrics across various clinical texts. This benchmark is structured around three sub-tasks: Sentence-level Human Rating, Paragraph-level Human Rating, and Comparison of Synthetic Reports, each targeting specific evaluation challenges. Both the RaTE-NER dataset and the RaTE-Eval benchmark have been made publicly available, aiming to foster the development of more effective evaluation metrics within the field of medical informatics.

Our extensive experiments demonstrate the superiority of **RaTEScore**. We initially tested our metric against others on the public dataset ReXVal (Yu et al., 2023a), and it shows superior alignment with human preference. Considering ReXVal’s limitation to chest X-ray reports, further testing was conducted on the diverse sub-tasks of RaTE-Eval, where RaTEScore consistently outperformed other metrics. We also conduct ablation studies to validate the effectiveness of different individual components in our pipeline.

## 2 Methods

In this section, we start by properly formulating the problem, and introducing the pipeline of our metric (Sec. 2.1). Then, we detail each of the modules for our metric computation, *i*.*e*., medical named entity recognition (Sec. 2.2), synonym disambiguation encoding (Sec. 2.3), and the final scoring procedure (Sec. 2.4). Lastly, we present the implementation details at each stage.

### 2.1 General Pipeline

Given two radiological reports, one is the ground truth for reference, denoting as *x*, and the other candidate for evaluation as 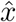. We aim to define a new similarity metric 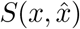, that enables comparison of two radiological reports at the entity level, thus better reflecting their clinical consistency.

As shown in Figure 2, our pipeline contains three major components: namely, a medical entity recognition module (Φ_NER_(•)), a synonym disambiguation encoding module (Φ_ENC_(•)), and a final scoring module (Φ_SIM_(•)). First, we extract the medicial entities from each piece of radiological text, then encode each entity into embeddings that are aware of medical synonym, formulated as:

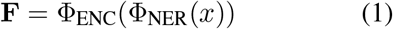

where **F** contains a set of entity embeddings. Similarly, we can get 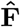 for 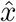. Then, we can calculate the final similarity on the entity embeddings as:

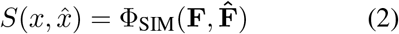

**Figure 2:**
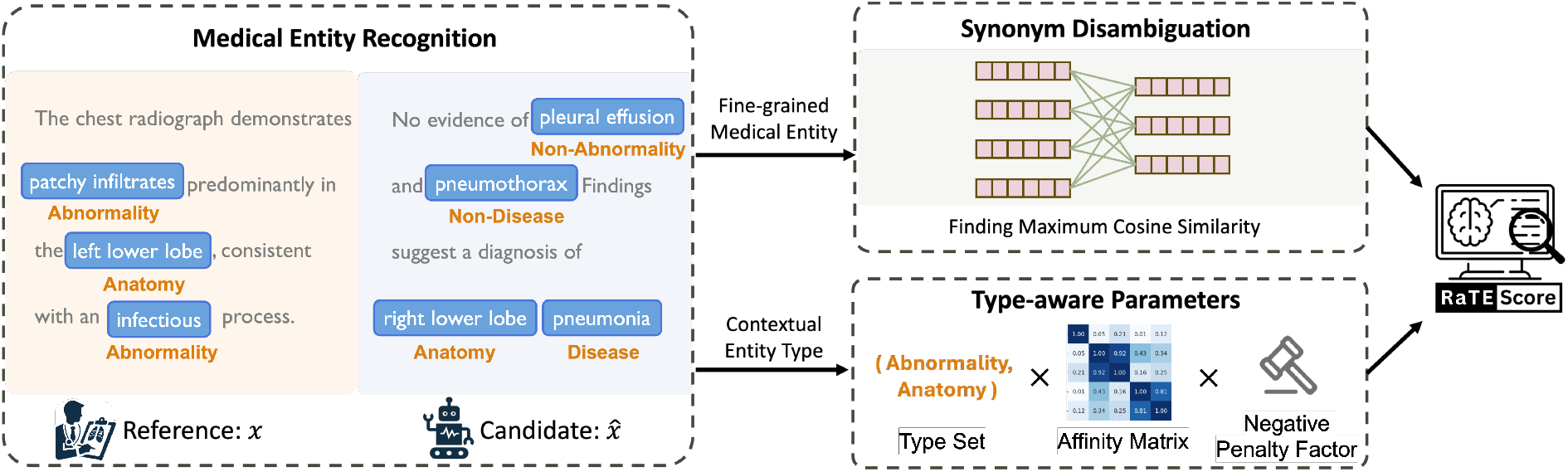
Illustration of the Computation of RaTEScore. Given a reference radiology report *x*, a candidate radiology report 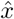, we first extract the medical entity and the corresponding entity type. Then, we compute the entity embedding and find the maximum cosine similarity. The RaTEScore is computed by the weighted similarity scores that consider the pairwise entity types.

In the following sections, we will detail each of the components.

### 2.2 Medical Named Entity Recognition

In the medical named entity recognition module, our goal is to decompose each radiological report by identifying a set of entities:

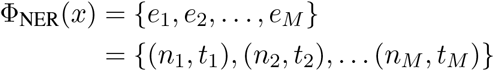

Similarly, we can also get 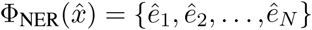, where *M, N* denote the total number of entities extracted from each text respectively. Each entity *e*_*i*_ is defined as a tuple (*n*_*i*_, *t*_*i*_), where *n*_*i*_ is the name of the entity and *t*_*i*_ denotes its corresponding type. For instance, the tuple (‘pneumonia’, ‘Disease’) represents the entity ‘pneumonia’ categorized under the entity type ‘Disease’.

Overall, we categorize the entity types into five distinct groups within radiological contexts: {*Anatomy, Abnormality, Disease, Non-Abnormality, Non-Disease*}. Specifically, ‘Abnormality’ refers to notable radiological features such as masses, effusion, and edema. Conversely, ‘Non-Abnormality’ denotes cases where such abnormalities are negated in the context, as illustrated by the classification of ‘pleural effusion’ in the statement ‘No evidence of pleural effusion’. Compared to ‘Abnormality’, ‘Disease’ in radiology reports are more high-level, mainly about final main professional diagnosis conclusions, terms such as ‘pneumonia’ or ‘lymphadenopathy’.

#### RaTE-NER Dataset

To support the development of our medical entity recognition module, we have constructed the **RaTE-NER** dataset, a largescale, radiological named entity recognition (NER) dataset, including 13,235 manually annotated sentences from 1,816 reports within the MIMIC-IV database, that spans 9 imaging modalities and 23 anatomical regions, ensuring comprehensive coverage. Given that reports in MIMIC-IV are more likely to cover common diseases, and may not well represent rarer conditions, we further enriched the dataset with 33,605 sentences from the 17,432 reports available on Radiopaedia (Rad), by leveraging GPT-4 and other medical knowledge libraries to capture intricacies and nuances of less common diseases and abnormalities. More details can be found in the Appendix A.2. We manually labeled 3,529 sentences to create a test set. As shown in Table 2 and Table 1, the **RaTE-NER** dataset offers a level of granularity not seen in previous datasets, with comprehensive entity annotations within sentences, that enables to train models for medical entity recognition within our analytical pipeline.

**Table 1:** RaTE-NER Dataset Statistics (Report-level): The number of sentences from medical reports of each data source.

| Data Source | Train Set | Dev Set | Test Set |
| --- | --- | --- | --- |
| MIMIC-IV | 10588 | 1323 | 1324 |
| Radiopaedia | 30005 | 3600 | 3529 |
| **Total Reports** | **40593** | **4923** | **4853** |

**Table 2:** RaTE-NER Dataset Statistics (Entity-level): The numbers outside and inside the brackets denote the total number of entities for each type, and the number of non-duplicate entities, respectively.

|  | MIMIC-IV |  |  |
| --- | --- | --- | --- |
|  | Train Set | Dev Set | Test Set |
| Anatomy | 9034 (4314) | 1188 (828) | 1140 (765) |
| Abnormality | 5579 (4047) | 760 (657) | 605 (513) |
| Non-Abnormality | 4182 (1528) | 479 (274) | 514 (253) |
| Disease | 1675 (1220) | 189 (169) | 178 (164) |
| Non-Disease | 3482 (965) | 424 (268) | 457 (264) |
|  | Radiopaedia |  |  |
|  | Train Set | Dev Set | Test Set |
| Anatomy | 34110 (14051) | 4145 (2629) | 4471 (2889) |
| Abnormality | 33863 (23352) | 4021 (3386) | 4265 (3365) |
| Non-Abnormality | 3878 (2280) | 473 (325) | 605 (420) |
| Disease | 9639 (7385) | 1118 (1044) | 741 (659) |
| Non-Disease | 2467 (1540) | 268 (220) | 183 (142) |
| **Total Entities** | **107909 (60682)** | **13065 (9800)** | **13159 (9434)** |

### 2.3 Synonym Disambiguation Encoding

To address the challenge from synonyms in the evaluation process, such as reconciling terms like “lung” and “pulmonary”, we propose to map each entity name into embedding space, where synonyms are positioned closely together, utilizing a medical entity encoding module trained with extensive medical knowledge. This module, represented as: *f*_*i*_ = Φ_ENC_(*n*_*i*_), with *f*_*i*_ denotes the vector embedding for the entity name. Consequently, we compile these into a set of entity embeddings: **F** = (*f*_1_, *t*_1_), (*f*_2_, *t*_2_),….}. A similar set, 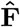, is constructed for the candidate text using the same methodology. For this encoding process, We adopt an off-shelf retrieval model, namely, BioLORD (Remy et al., 2024), which is trained specifically on medical entity-definition pairs and has proven effective in measuring entity similarity.

### 2.4 Scoring Procedure

Upon obtaining the encoded entity set from each decomposed radiological report, we proceed to the final scoring procedure. We first define the similarity metric between a candidate entity and a reference report, that is established by selecting an entity from the referenced text based on the cosine similarity of their name embeddings:

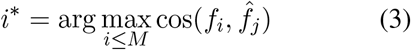

where cos 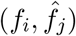 measures the cosine similarity between two entity name embeddings. The entity *e*_*i*_*, which best matches *ê*_*j*_ from the reference text, is chosen for further comparison. The overall similarity score, 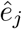, is then computed as follows:

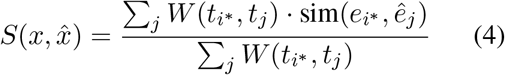

Here, *W* is a learnable 5 × 5 affinity matrix between the five entity types, where *W* (*t*_*i*_, *t*_*j*_) represents an element of the matrix, and sim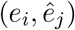 is an entitywise similarity function, defined as:

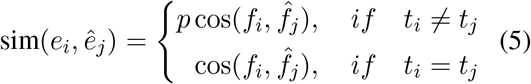

where we generally follow the cosine similarity on the name embedding, with a learnable penalty value *p* to punish the type mismatch. For example, when comparing entities with identical names but different types—such as (‘pleural effusion’, ‘Abnormality’) and (‘pleural effusion’, ‘Non-Abnormality’)—the penalty term *p* is applied to adjust the similarity score appropriately.

Additionally, the similarity between different entity types may be weighted differently in medical scenarios due to their clinical significance. For example, the similarity between two ‘Abnormality’ entities is of much greater importance than the similarity between two ‘Non-abnormality’ entities. This is because all body parts are assumed to be normal in radiology reports by default, and minor expression errors in normal findings will not critically impact the report’s correctness. Therefore, we introduce *W* to account for this clinical relevance.

Finally, due to the order of performing max indexing for selecting referenced entities and weighted sum pooling on all candidate entities, the final similarity metric 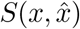 does not comply with the commutative law. 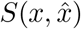 and 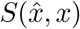 can be analogous to precision and recall respectively. Thus, our final **RaTEScore** is defined as the harmonic mean of 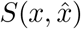 and 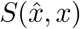, following classical F_1_-score format:

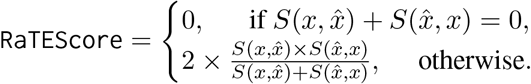

### 2.5 Implementation Details

In this section, we describe the implementation details for the three key modules. *First*, for medical named entity recognition, we train a BERT-liked model on **RaTE-NER** dataset with two main-stream NER architectures, namely, Span-based and IOB-based models. For the Span-based method, we follow the setting of PURE (the Princeton University Relation Extraction system) entity model (Zhong and Chen, 2021) and for the IOB-based method, we follow DeBERTa v3 (He et al., 2022, 2020). We show more detailed implementation parameters for the two training schemes in Appendix A.9. Additionally, we also try to initialize the NER model with different pre-trained BERT. More comparison of the two training schemes and different BERT initializations will be present in the ablation study. *Second*, For the synonym disambiguation encoding, we directly use the off-shelf BioLORD-2023-C model version. Ablation studies are also conducted in Section 4. *Third*, for the final scoring module, we learn the affinity matrix *W* and negative penalty factor *p* leveraging TPE (Tree-structured Parzen Estimator) (Bergstra et al., 2011) with a small set of human rating data.

## 3. RaTE-Eval Benchmark

To effectively measure the alignment between automatic evaluation metrics and radiologists’ assessments in medical text generation tasks, we have established a comprehensive benchmark, **RaTE-Eval**, that encompasses three tasks, each with its official test set for fair comparison, as detailed below. The comparison of RaTE-Eval Benchmark and existed radiology report evaluation Benchmark is listed in Table 3.

**Table 3:**
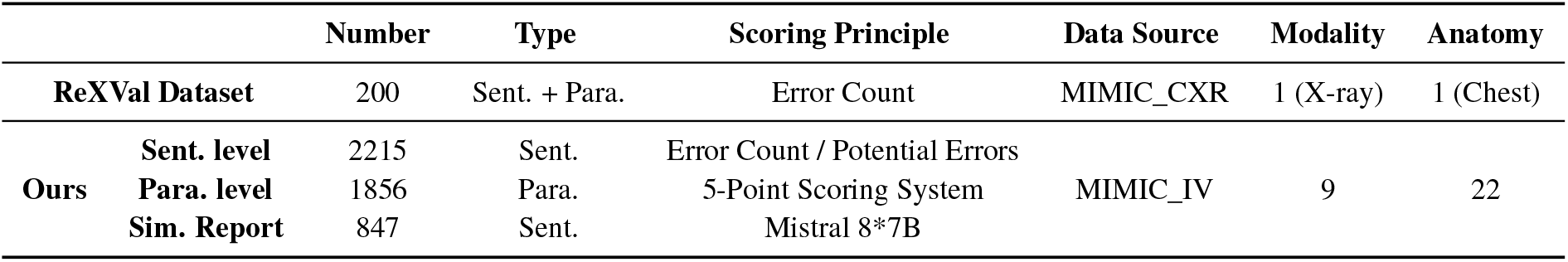
Comparison of RaTE-Eval Benchmark and existed radiology report evaluation Benchmark.

### Sentences-level Human Rating

Existing studies have predominantly utilized the ReXVal dataset (Yu et al., 2023b), which requires radiologist annotators to identify and count errors in various potential categories. The metric’s quality is assessed by the Kendall correlation coefficient between the total number of errors and the result from the automatic metric. The possible error categories are as follows:

- False prediction of finding;
- Omission of finding;
- Incorrect location/position of finding;
- Incorrect severity of finding;
- Mention the comparison that is absent from the reference impression;
- Omission of comparison describing a change from a previous study.

Building on this framework, we introduce two improvements to enhance the robustness and applicability of our benchmark: **(1) normalization of error counts**: recognizing that a simple count of errors may not fairly reflect the informational content in sentences, we have adapted the scoring to annotate **the number of potential errors**. Specifically, we computed the sum of both correct and incorrect findings in the reference sentence. This sum represents the total number of potential errors that could occur. This approach normalizes the counts, ensuring a more balanced assessment across varying report complexities. **(2) diversification of medical texts**: unlike existing benchmarks that are limited to Chest x-rays from the MIMIC-CXR dataset (Johnson et al., 2019), our dataset includes **2215** reports spanning **9** imaging modalities and **22** anatomies from the MIMIC-IV dataset (Johnson et al., 2020), the involving imaging modalities and anatomies are listed in Appendix A.3.

Specifically, our annotation process is as follows: First, we divide the MIMIC-IV dataset into 49 subsets based on modality and anatomy. To conduct sentence-level evaluation, we split the report paragraphs into individual sentences by periods and remove the duplicates. Next, we randomly sample 25 sentences from each subset to create a reference report list and sample another 1000 reports to form a candidate report list. Subsequently, we use several evaluation metrics—BLEU, ROUGE, BERTScore, CIDEr, and our proposed RaTEScore to identify the most similar one from the candidate report list for each report in the reference report list. We take the union of all the metric results as the report pairs for manual annotations. Finally, each case in the annotation reports was annotated by two experienced radiologists with over five years of clinical practice. For each candidate report and the corresponding reference report, they are required to count errors in six provided categories as well as the number of potential errors, where the error refers to the candidate report’s error based on the reference report.

The final human rating result is computed by dividing the total error count by the the number of potential errors. The final sentence-level benchmark is composed of 2215 reference report sentences, candidate report sentences and their rating score. For parameter search (Sec. 2.5), we divided all reports into a training set and a test set at an 8:2 ratio, to identify the most effective parameters that align with human scoring rules.

### Paragraph-level Human Rating

Given that medical imaging interpretation commonly involves the evaluation of lengthy texts rather than isolated sentences, we also incorporate paragraph-level assessments into our analysis, from the MIMIC-IV reports. However, as it is challenging for humans to completely count all errors in long paragraphs accurately, we established a 5-point scoring system for our evaluations, following the RadPEER (Goldberg-Stein et al., 2017) - an internationally recognized standard for radiologic peer review. They defines “Concur with Interpretation” as “correct diagnosis”. In our annotation process, due to the difficulty of counting errors in paragraph-level reports (too detailed and some errors are not of great clincial significance), we instructed the radiologists to approximate the ratio of correct diagnosess based on their clinical judgement, following the ‘RadPEER’ standard, which is more aligned with the human rating system for report writing in clinical.

The scores range from 5, denoting a perfectly accurate report, to 0, that indicates the report without any correct observations. Detailed scoring criteria are provided in Appendix A.4, guiding radiologists on how to assign scores at different levels.

Specifically, our annotation process is as follows: first, we divide the MIMIC-IV dataset into 49 subsets based on modality and anatomy. Next, we sample 20 reports from each subset to create a reference list and 500 reports to form a candidate list. The report selection process is the same as sentence-level human rating. For each candidate report and the corresponding reference report, the radiologists are required to give a 5-point score.

The final benchmark in paragraph-level is composed of 1856 reference reports, candidate reports and their rating score. Similarly, for parameter search (Sec. 2.5), we also divide all reports into training set and a test set at an 8:2 ratio.

### Rating on the Synthetic Reports

Here, we aim to evaluate the sensitivity of our metric on handling synonyms and negations using synthetic data. Specifically, we employed Mixtral 8×7B (Jiang et al., 2024), a sophisticated open-source Large Language Model (LLM), to rewrite **847** reports from the MIMIC-IV dataset. The rewriting was guided by two tailored prompts:

*You are a specialist in medical report writing, please rewrite the sentence, you can potentially change the entities into synonyms, but please keep the meaning unchanged*.

On the other hand, opposite reports were generated with:

*You are a specialist in medical report writing, please rewrite the following medical report to express the opposite meaning*.

This process results in a test set comprising triads of reports: the original, a synonymous version, and an anonymous version, detailed further in Appendix A.5. Ideally, effective evaluation metrics should demonstrate higher scores for synonymous reports compared to anonymous reports, thereby more accurately reflecting the true semantic content of the reports.

## 4 Experiments

In this section, we start by introducing the baseline evaluation metrics. Later, we compare the different metrics with our proposed RaTEScore on both ReXVal and RaTE-Eval benchmarks. Lastly, we present details for the ablation studies.

### 4.1 Baselines

We use the following metrics as baseline comparisons: BLEU (Papineni et al., 2002), ROUGE (Lin, 2004), METEOR (Banerjee and Lavie, 2005), CheXbert (Smit et al., 2020; Yu et al., 2023a), BERTScore (Zhang et al., 2020), SPICE (Anderson et al., 2016) and RadGraph F1 (Yu et al., 2023a). Detailed explanations of these metrics can be found in the Appendix A.6.

### 4.2 Results in ReXVal dataset

Our initial evaluation adopts the public ReXVal dataset, where we calculated the Kendall correlation coefficient to measure the agreement between our RaTEScore and the average number of errors identified by six radiologists. Our analysis was conducted under identical conditions to those of baseline methods. Given that the reports in ReXVal vary significantly in length, predominantly featuring longer documents, we applied a type weight matrix with parameters specifically fine-tuned on our long-report benchmark training set. As detailed in Table 4, RaTEScore demonstrated a Kendall correlation coefficient of 0.527 with the error counts, surpassing all existing metrics.

**Table 4:** Results in ReXVal dataset: * denotes the result report in (Yu et al., 2023a).

|  | RadGraph F1 | BERTScore | CheXbert | BLEU | Ours |
| --- | --- | --- | --- | --- | --- |
| Kendall $\tau$ | 0.515* | 0.511* | 0.499* | 0.462* | **0.527** |

While further examining instances with notable deviations in Appendix A.7, a primary observation was that ReXVal’s protocol tends to count six types of errors uniformly, without accounting for variations in report length. This approach leads to discrepancies where a single-sentence report with one error type and a twenty-sentence report with the same error count receive equivalent scores. To address this issue, our **RaTE-Eval** benchmark can be better suited to distinguish such variations, by normalising the total error counts with potential error counts.

### 4.3 Results in RaTE-Eval benchmark

#### On Sentence-level Rating

As illustrated in Fig. 3, our model achieved a Pearson correlation coefficient of 0.54 on the RaTE-Eval short sentence benchmark, significantly outperforming the existing baselines. These results underscore the inadequacy of methods that predominantly rely on term overlap for evaluations within a medical context. While entity-based metrics like RadGraph F1 show notable improvements, they still do not reach the desired level of efficacy on an extensive benchmark encompassing multi-modal, multi-region reports. This shortfall largely attributes to the limited scope of training vocabulary in these methods.

**Figure 3:**
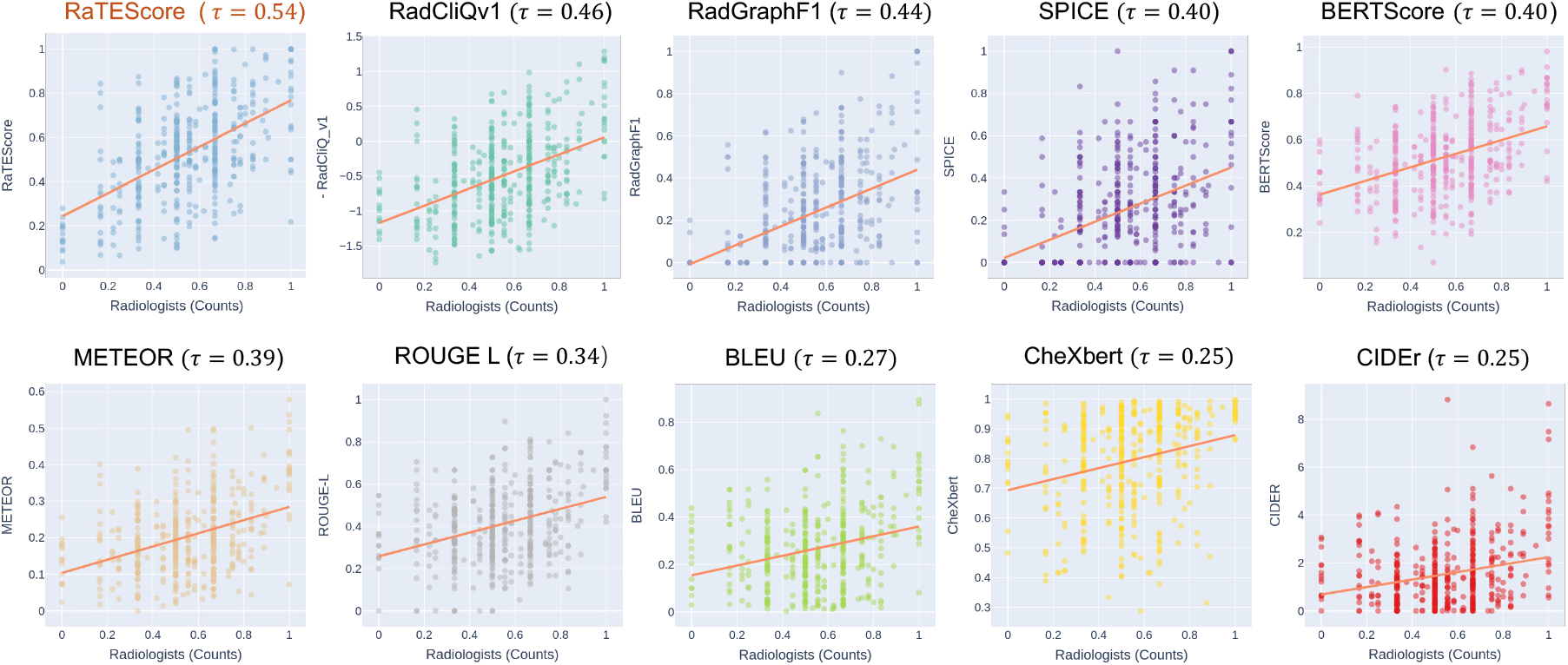
Results in RaTE-Eval Benchmark: Correlation Coefficients with Radiologists Results (sentence-level). our metric exhibits the highest Pearson correlation coefficient with the radiologists’ scoring. Note that the scores on the horizontal axis are experts counting various types of errors normalized by the potential error types that could occur in the given sentence, and subtracting this normalized score from 1 to achieve a positive correlation.

#### On Paragraph-level Rating

From the results in Table 5, it can be observed that **RaTEScore** shows a significantly higher correlation with radiology experts compared to other existing metrics, across various measures of correlation. Metrics that focus on identifying key entities, such as RadGraph F1, SPICE, and ours, consistently demonstrate stronger correlations than those reliant on mere word overlap, thereby supporting our primary assertion that critical statements in medical reports are paramount. Furthermore, metrics that accommodate synonyms, such as METEOR, outperform those that do not, such as BLEU and ROUGE. Significantly, **RaTEScore** benefits from a robust NER model trained on our comprehensive dataset, **RaTE-NER**, which spans multiple modalities and anatomical regions, not just Chest x-rays, resulting in markedly higher correlations.

**Table 5:** Results in RaTE-Eval Benchmark: Correlation coefficients with radiologists and accuracy for whether the synonym sentence can achieve higher scores than the antonymous one on Synthetic Reports.

|  | Paragraph-level Correlations |  |  | Simulations |
| --- | --- | --- | --- | --- |
| | Pearson $\tau$ | Kendall $\tau$ | Spearman $\tau$ | Acc |
| RadGraph | 0.624 | 0.439 | 0.582 | 0.463 |
| BERTScore | 0.599 | 0.413 | 0.555 | 0.140 |
| CheXbert | 0.496 | 0.294 | 0.403 | 0.666 |
| BLEU | 0.409 | 0.289 | 0.404 | 0.119 |
| ROUGE_L | 0.572 | 0.396 | 0.567 | 0.117 |
| SPICE | 0.623 | 0.453 | 0.605 | 0.140 |
| METEOR | 0.599 | 0.422 | 0.567 | 0.168 |
| **Ours** | **0.653** | **0.462** | **0.608** | **0.670** |

#### Results on Synthetic Reports

To further showcase the effectiveness of our proposed **RaTEScore**, we examined its performance on the synthetic test set. This dataset, being synthesized, allows us to use accuracy (ACC) as a measure to evaluate performance. Specifically, we assess whether the synonymously simulated sentences received higher scores than their antonymous counterparts. The results, presented in Table 5, demonstrate that our model excels in managing synonym and antonym challenges, affirming its robustness in nuanced language processing within a medical context.

### 4.4 Ablation Study

In this ablation section, we investigate the pipeline from two aspects: namely, the design of NER model, the effect of different off-the-shelf synonym disambiguation encoding module.

#### 4.4.1 NER Module Discussion

Here, we discuss the performance of our NER module in three parts: training schemes, initialization models, and data composition.

##### Training Schemes

To select the most suitable NER model for training, we compare IOB-based and Span-based NER training schemes on the whole RaTE-NER test set. As shown in Table 6, the IOB scheme overall extracts more comprehensive entities, but the recall is lower against the Spanbased approach.

**Table 6:** Ablation Study on NER Model Schemes.

|  | Initialized BERT | Pre | Recall | F1 | Acc |
| --- | --- | --- | --- | --- | --- |
| **IOB.** | DeBERTa_v3 | **0.567** | 0.575 | 0.571 | 0.754 |
|  | Medical-NER | 0.559 | 0.572 | 0.565 | **0.759** |
| **Span.** | BiomedBERT | 0.556 | 0.676 | **0.610** | 0.730 |
|  | SapBERT | 0.560 | 0.658 | 0.605 | 0.731 |
|  | BlueBERT | 0.554 | 0.657 | 0.601 | 0.726 |
|  | MedCPT-Q-Enc. | 0.470 | **0.682** | 0.556 | 0.678 |
|  | BioLORD-2023-C | 0.555 | 0.664 | 0.605 | 0.727 |

##### Initialization Models

Additionally, as shown in Table 6, we also try a sequential pre-trained BERT model for initialization, *i*.*e*., DeBERTa_v3 (He et al., 2022), Medical-NER (Clinical-AI-Apollo, 2023), BioMedBERT (Chakraborty et al., 2020), BlueBERT (Peng et al., 2019), MedCPT-Q-Enc. (Jin et al., 2023), and BioLORD-2023-C (Remy et al., 2024). Detailed description for each model can be found in Appendix A.8. We apply various models in different training schemes based on their pre-training tasks. For example, Medical-NER is pre-trained with IOB-based NER tasks on other tasks thus we still finetune it in the same setting. Comparing Medical-NER and DeBERTa_v3, pretraining on other NER datasets does not improve much. Different types of BERT also perform fairly for the Span-based method. Based on the results, our final scores are all based on the IOB scheme with DeBERTa_v3.

##### Data Ablation

Our RaTE-NER data is composed of two distinct parts, and we conducted experiments to highlight the necessity of both. As shown in Table 7, ‘R.’ denotes data from Radiopaedia, while ‘M.’ refers to the data from MIMIC-IV. By combining these two parts (denoted as ‘R.+M.’), we observe a significant improvement in the final NER performance, with an increase of 0.030 in F1 and 0.010 in ACC. This underscores the importance of incorporating each dataset component.

**Table 7:** Ablation Study on NER Training Data. R. denotes data from Radiopaedia and M. denotes data from MIMIC-IV.

| Training Data | Pre | Recall | F1 | Acc |
| --- | --- | --- | --- | --- |
| R. | 0.525 | 0.558 | 0.541 | 0.727 |
| M. | 0.515 | 0.550 | 0.531 | 0.744 |
| **R. + M.** | **0.567** | **0.575** | **0.571** | **0.754** |

#### 4.4.2 Entity Encoding Module Discussion

For the evaluation of Entity Encoding Module, we compare several off-the-shelf entity encoding models trained using different approaches on the sentence-level correlation task of RaTE-Eval. BioLORD-2023-C (Remy et al., 2024) is trained on medical entity concepts, MedCPT-Query-Encoder (Jin et al., 2023) is trained on PubMed user click search logs, while RadBERT (Chambon et al., 2023), CXR-BERT (Boecking et al., 2022), and BioViL-T (Bannur et al., 2023) are pre-trained on a large corpus of radiology texts. As shown in Table 8, BioLORD, due to its original training goal covering medical entity normalization, which aligns with our needs in the Entity Encoding module, achieved the best performance. Based on these results, we selected BioLORD-2023-C as the base model for our Entity Encoding Module.

**Table 8:**
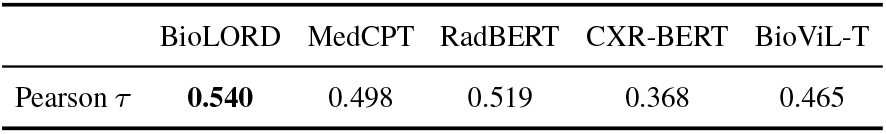
Ablation Study on Pretrained Model of Entity Encoding Module.

## 5 Related Work

### 5.1 General Text Evaluation Metric

Automated scoring methods allow for a fair evaluation of the quality of generated text. BLEU (Papineni et al., 2002), ROUGE (Lin, 2004) was originally designed for machine translation tasks, focusing on word-level accuracy. METEOR (Banerjee and Lavie, 2005) adopts a similar design, taking into account synonym matching and word order. SPICE (Anderson et al., 2016) uses the key objects, attributes, and their relationships to compute the metric. BERTScore (Zhang et al., 2020), a modelbased method, assigns scores to individual words and averages these scores to evaluate the text’s overall quality, facilitating a more detailed analysis of each word’s contribution.

### 5.2 Radiological Text Evaluation Metric

With the advancement of medical imaging analysis, researchers have recognized the importance of evaluating the quality of radiology text generation. Metrics such as CheXbert F1 (Smit et al., 2020) and RadGraph F1 (Yu et al., 2023a) are based on medical entity extraction models. However, CheXbert can only annotate 14 chest abnormalities, and RadGraph F1 (Jain et al.) is only trained on chest X-ray modality. MEDCON (Yim et al., 2023) expands the extraction range by QuickUMLS package (Soldaini and Goharian, 2016), which relies on a string match algorithm that is not flexible. RadCliQ (Yu et al., 2023a) performs ensembling with BLEU, BERTScore, CheXbert vector similarity, and RadGraph F1 for a comprehensive yet less interpretable evaluation. These metrics calculate the overlap between reference and candidate sentences while overlooking the issue of synonymy. Recently, metrics using Large Language Models (LLMs) such as GPT-4, such as G-Eval (Liu et al., 2023), LLM-as-a-Judge (Zheng et al., 2024), and LLM-RadJudge (Wang et al., 2024) have emerged, closely mimic human evaluation levels. However, these methods are unexplainable and may have potential subjective bias. Besides, their high computational cost also limits them for statistic robust large-scale evaluation.

### 5.3 Medical Named-Entity Recognition

The MedNER task targets extracting medical-related entities from given contexts. Great efforts have been made in this domain (Jin et al., 2023; Monajatipoor et al., 2024; Keloth et al., 2024; Li and Zhang, 2023; Chen et al., 2023). Inspired by the success of this work, we believe MedNER models are strong enough to simplify and structure complex clinical texts, thus reducing the difficulty of automatically comparing two clinical texts. The most related work to ours is RadGraph (Jain et al.) which trained an NER model for Chest X-ray reports while we are targeting more general clinical reports regardless of their type.

## 6 Conclusion

In this work, we propose a new lightweight, explainable medical free-form text evaluation metric, **RaTEScore**, by comparing two medical reports on the entity level. In detail, first, we build up a new medical NER dataset, **RaTE-NER** targeting a wide range of radiological report types and train a NER model on it. Then, we adopt this model to simplify the complex radiological reports and compare them on the entity embedding level leveraging an extra synonyms disambiguation encoding model. Our final RaTEScore correlates strongly with clinicians’ true preferences, significantly outperforming previous metrics both on the former existing benchmark and our new proposed **RaTE-Eval**, while maintaining computational efficiency and interpretability.

## Data Availability

All data produced in the present work are contained in the manuscript.

## Limitations

Although our proposed metric, RaTEScore, has performed well across various datasets, there are still some limitations. First, in the synonym disambiguation module, we evaluated the performance of several existing models and directly ultilized them without fine-tuning specifically for the evaluation scenario, which could be enhanced in the future. Furthermore, while we expanded from single-modality radiological report evaluation to multimodal whole-body imaging, we still only considered the issues within the radiological report scenario and did not extend to other medical contexts beyond radiology, nor to the evaluation of other medical tasks, like medical QA, summarization task. These areas require more exploration.

## Acknowledgements

This work is supported by the National Key R&D Program of China (No. 2022ZD0160702).

## A Appendix

### A.1 Scoring Example

In this section, we will show an example of calculating RaTEScore. Given a radiology report pair:

**Referenced** *x*: A Foley catheter is in situ.

**Candidate** 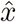: A Foley catheter is not in place.

For simplicity, we will only describe the calculation procedure for 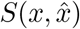 in text, and the calculation procedure for 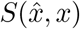 is similar.

We first conduct **Medical Named Entity Recognition** to decompose the natural text into entities. For the referenced report, the entities list is: {(“Foley catheter”, Anatomy), (“in situ”, Non-Abnormality) } and for the candidate report is {(“Foley catheter”, Anatomy), (“not in place”, Abnormality) }. Subsequently, these extracted entities are processed through the **Synonym Disambiguation Encoding Module**, which encodes the “Foley catheter” and “in situ” into feature embedding. Finally, during the **Scoring Procedure**, we pick out the most similar entity in the referenced report for each entity in the candidate report, *i*.*e*., “Foley catheter” paired with “Foley catheter” in the reference, and “not in place” with “in situ”. Then, we get two cosine similarity scores based on the text embedding, 1.0 for “Foley catheter” and 0.83 for “not in place”. The similarity score between (“not in place”, Abnormality) and (“in situ”, Non-Abnormality) will be further multiplied with a penalty factor *p* as 0.37 while the other similarity is maintained since they have the same entity type. At Last, we calculate the weighted combination of the two. The weights are derived from a learnable attribution matrix *W* corresponding to these type combinations, as 0.91 and 0.94 respectively. The calculation formulation is as follows:

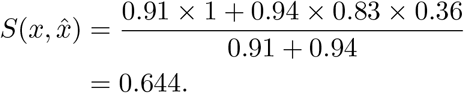

Similarly, we can get the other similarity:

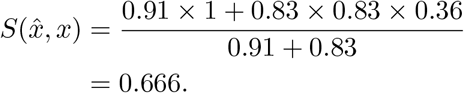

Notably, the only difference between the two similarity scores in this case lies in the weight between (“in situ”, Non-Abnormality) and (“not in place”, Abnormality). Due to the comparison directions, in 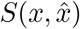, *W* (Non-Abnormality, Abnormality) as 0.94 is adopted and in the other hand, *W* (Abnormality, Non-Abnormality) as 0.83 is adopted. The final score is computed as follows:

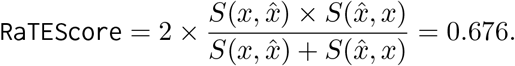

#### A.2 Automatic Annotation Approach

Here, we introduce our automatic approach to construct a part of our **RaTE-NER** dataset, sourced from 19,263 original reports obtained from Radiopaedia (Rad) and covering 9 modalities and 11 anatomies. As shown in Figure 4, leveraging the latest LLM GPT-4 combined with other comprehensive medical knowledge bases, we develop a new automated medical NER and relation extraction dataset construction pipeline.

**Figure 4:**
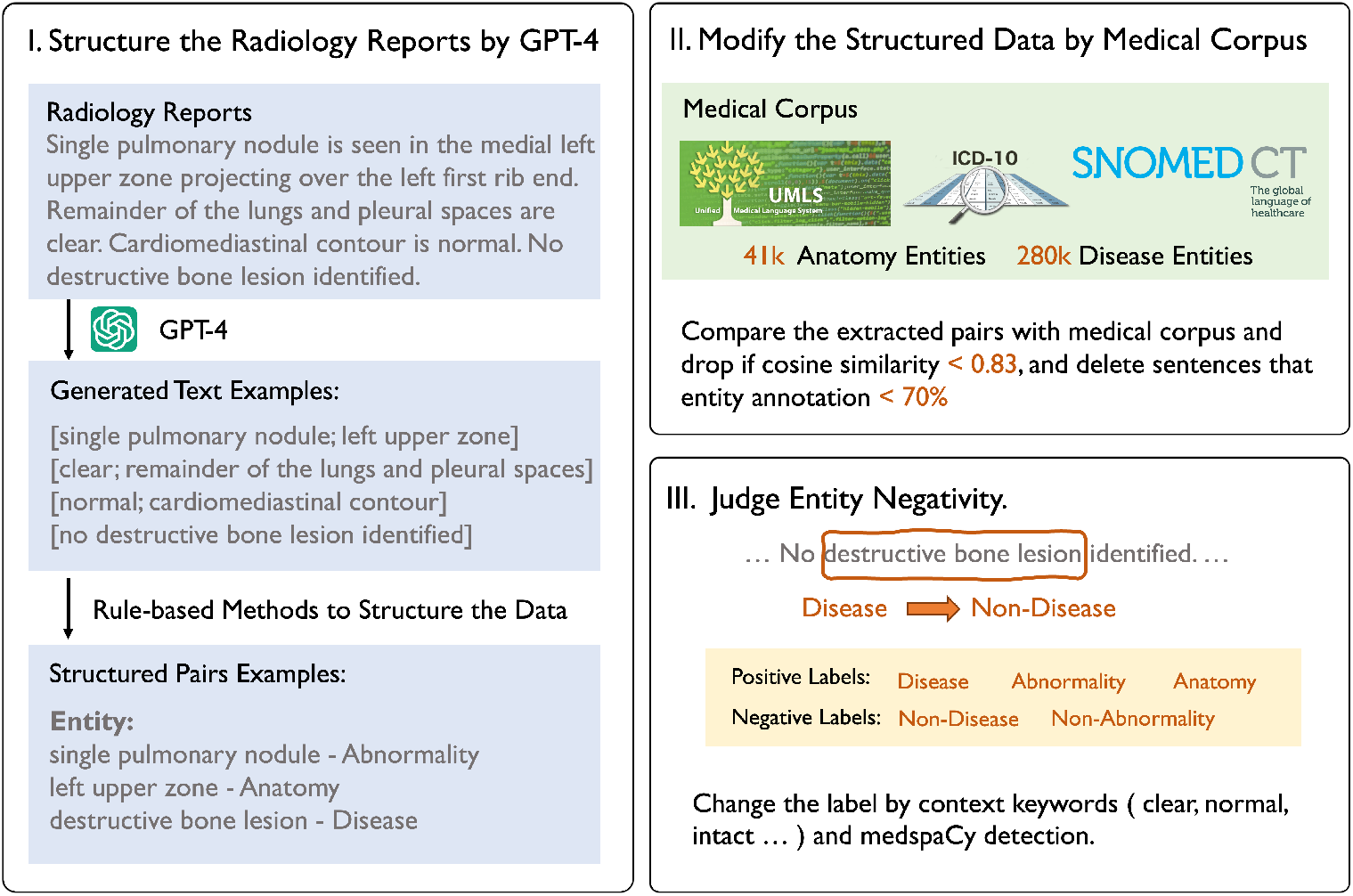
Data Curation Procedure.

Specifically, we manually annotate several reports at the required granularity and adopt few-shot prompts with GPT-4 to initially establish an NER dataset.

**GPT-4 prompt**:

*You are an AI assistant specializing in radiology reports reading. You are provided with a medical caption. Extract the entities and decide their type from organ, abnormal description or disease. Collect the organ and description together if the description modifies the organ. Leave disease alone. Make sure that the description is about the abnormality but not position*.

*The output should follow this format: [organs; abnormal description] or [disease]. All words in [] should belong to the original sentence*.

**Few-shot examples**:

‘context’: *“The sentence is: Hetergeneous and nodular enhancement of the liver with pre-contrast HU of -4 (!) indicating hepatic steatosis*.*”*,

‘response’: *“ [ liver; Hetergeneous and nodular enhancement ] [ liver; pre-contrast HU of -4 ] [ hepatic steatosis ] “*

Following this, we build a robust medical entity library, integrating UMLS (Bodenreider, 2004), Snomed CT (Donnelly et al., 2006), ICD-10 (ICD), and other knowledge bases, then, compare all extracted entities using the MedCPT (Jin et al., 2023) model for similarity. During the comparison process, entities with cosine similarity lower than 0.83 were filtered out. Most entities below this threshold did not meet our requirements. Subsequently, we removed sentences with an entity annotation density lower than 0.7 at the sentence level. Finally, we use medspaCy (Eyre et al., 2021) and also key negative words detection in reports, such as “no”, “without”, “unremarkable”, “intact”, to determine the positive or negative polarity of each word in the sentence.

#### A.3 Involving Anatomies and Modalities in MIMIC-IV Data

In this section, we detail the imaging modalities and anatomies involved in MIMIC-IV dataset.

**Anatomy List:** NECK, TEETH, BRAIN, HEAD, CHEST, PELVIS, ABDOMEN, CARDIAC, HEAD-NECK, SOFT TISSUE, UP-EXT, OB, EXT, HIP, BREAST, SPINE, MAMMO, BRAIN-FACE-NECK, LOW-EXT, BONE, VAS-CULAR, BLADDER.

**Modality List:** CT, CTA, Fluoroscopy, Mammography, MRA, MRI, MRV, Ultrasound, X-Ray.

#### A.4 Guidelines for Radiologists

Referencing RadPEER (Goldberg-Stein et al., 2017), we set up a five-point scoring criteria, as shown in Table 9. During the annotation process, each report is compensated with $1 per report, with five reference reports separately.

**Table 9:** 5-point scoring system For Radiologists to Rate in Paragraph-level Human Rating of RaTE-Eval Benchmark.

| Score | Meaning | Explanation |
| --- | --- | --- |
| 5 | Correct | Most of the diagnosis results are correct. Most descriptions are the same. Some wrong description unlikely to be clinically significant. |
| 4 | Almost Correct | 75% of the diagnosis results are correct. Most descriptions are the same. Some wrong description likely to be clinically significant. |
| 3 | Partly Correct | 50% of the diagnosis results are correct. |
| 2 | Partly Incorrect | 25% of the diagnosis results are correct |
| 1 | Major Errors Present | Incorrect diagnosis. Maybe some negative descriptions are the same. |
| 0 | Total Different | No overlap for the described information. |

#### A.5 Example for Simulation Reports

In this section, we give an example for the simulation report generation:

**GT:** The appendix is well visualized and air-filled.

**REWRITE:** The appendix is seen and contains gas.

**OPPOSITE:** The appendix is poorly visualized and not air-filled.

#### A.6 Baselines

Herein, we will introduce the considered baselines:

- BLEU (Papineni et al., 2002) measures the precision of generated text by comparing ngram overlap between the generated report and reference reports.
- ROUGE (Lin, 2004) focuses on the recall of generated text by measuring the overlap of n-grams, similar to BLEU.
- METEOR (Banerjee and Lavie, 2005) combines precision, recall, and a penalty for fragmented alignments, while also considering words order and synonyms through WordNet (Fellbaum, 2010).
- CheXbert (Smit et al., 2020; Yu et al., 2023a) computes the cosine similarity between CheXbert model embedding of the reference report and candidate report.
- BERTScore (Zhang et al., 2020) utilizes a pretrained BERT model to calculate the similarity of word embeddings between candidate and reference texts.
- SPICE (Anderson et al., 2016) extracts key objects, attributes, and their relationships from descriptions to build a scene graph, and compares the two texts on the scene graph level.
- RadGraph F1 (Yu et al., 2023a) extracts the radiology entities and relations for Chest X-ray modality and computes the F1 score on the entity level.

#### A.7 Failure Cases in ReXVal Dataset

In this section, in order to better demonstrate the drawbacks of ReXVal dataset, we will give a failure case where two reports with different entity-wise errors while achieve the same scores.

**Report Pair 1:**

**GT:** ET tube within 1 cm of the carina. This was discussed with Dr. at 4 p.m. on by Dr. at time of interpretation. **Pred:** ET tube terminates approximately 3. 5 cm from the carina.

**Total Errors:** 1.33

**Report Pair 2:**

**GT:** In comparison with the study of xxx, there is again enlargement of the cardiac silhouette with elevation of pulmonary venous pressure. Opacification at the right base again is consistent with collapse of the right middle and lower lobes RECOMMEN-DATION(S): The tip of the right IJ catheter is in the mid to lower SVC.

**Pred:** In comparison with the study xxx, there is little change in the appearance of the monitoring and support devices. Continued substantial enlargement of the cardiac silhouette with relatively mild elevation of pulmonary venous pressure. Opacification at the right base silhouettes the hemidiaphragm and is consistent with collapse of the right middle and lower lobes.

**Total Errors:** 1.33

As shown in the examples, case 1 with only two entity errors scores 1.3, and the report that describes more than ten different entity errors also scores 1.3. Moreover, reports length less than 10 words commonly has zero errors in ReXVal, whereas reports longer than 25 words had an average error count greater than 3, simply because the texts are longer and may contain more potential errors. Therefore, ignoring normalization and directly using absolute error counting numbers as the score like ReXVal may present severe bias that longer sentences scoring lower and shorter sentences scoring higher.

#### A.8 Pretrained BERT Model Introduction

In this section, we will introduce our considered pre-trained BERT models in detail:

- DeBERTa_v3 (He et al., 2022) is an advanced version of the DeBERTa (He et al., 2020) model, which improves upon the BERT and RoBERTa models by incorporating disentangled attention mechanisms, enhancing performance on a wide range of natural language processing tasks.
- Medical-NER (Clinical-AI-Apollo, 2023) is a fine-tuned version of DeBERTa to recognize 41 medical entities. The specific training data is not publicly available.
- BioMedBERT (Chakraborty et al., 2020) previously named “PubMedBERT”, pretrained from scratch using abstracts and full-text articles from PubMed (Canese and Weis, 2013).
- BlueBERT (Peng et al., 2019) is a BERT model pre-trained on PubMed abstracts and clinical notes (MIMIC-III) (Johnson et al., 2016).
- MedCPT-Q-Enc. (Jin et al., 2023) is pretrained by 255M query-article pairs from PubMed search logs, and achieve SOTA performance on several zero-shot biomedical IR datasets.
- BioLORD-2023-C (Remy et al., 2024) is based on a sentence-transformers model and further finetuned on the entity-concept pairs.

#### A.9 NER Module Implementation Details

In the Medical Named Entity Recognition Module training scheme, we train the model on one NVIDIA GeForce GTX 3090 GPU with a batch size of 96 for 10 epochs while adopt different learning rates for different training schemes. For the Span-based method, we follow the setting of PURE entity model (Zhong and Chen, 2021), which uses a pre-trained BERT model to obtain contextualized representations and then fed into a feedforward network to predict the probability distribution of the entity. It combines a BERT (Devlin et al., 2019) model and a 3-layer MLP with head hidden dimension of 3096 for span classification. The span max length is 8. In the training stage, we set the learning rate as 6e-6. For the IOB-based method, each token is labeled as ‘B-’ (beginning of an entity), ‘I-’ (inside an entity), or ‘O’ (outside of any entity). We directly fine-tune the pre-trained BERT to perform a token classification task. Specifically, we add a linear layer to the output embedding of a BERT-liked model, which is fine-tuned utilizing a corpus of annotated entity data to predict the entity label for each token. We use a learning rate of 1e-5 for the IOB-based training scheme.

## References

ICD-10-CM. https://www.icd10data.com/ICD10CM/Codes. Accessed: Dec.2023.

Radiopaedia.org. https://radiopaedia.org. Accessed: May 2023.

Jean-Baptiste Alayrac, Jeff Donahue, Pauline Luc, Antoine Miech, Iain Barr, Yana Hasson, Karel Lenc, Arthur Mensch, Katherine Millican, Malcolm Reynolds, et al. 2022. Flamingo: a visual language model for few-shot learning. Advances in Neural Information Processing Systems, 35:23716–23736.

Peter Anderson, Basura Fernando, Mark Johnson, and Stephen Gould. 2016. Spice: Semantic propositional image caption evaluation. In Proceedings of European Conference on Computer Vision (ECCV), pages 382–398.

Rohan Anil, Andrew M Dai, Orhan Firat, Melvin Johnson, Dmitry Lepikhin, Alexandre Passos, Siamak Shakeri, Emanuel Taropa, Paige Bailey, Zhifeng Chen, et al. 2023. Palm 2 technical report. arXiv preprint arXiv:2305.10403.

Satanjeev Banerjee and Alon Lavie. 2005. Meteor: An automatic metric for mt evaluation with improved correlation with human judgments. In Proceedings of the Acl Workshop on Intrinsic and Extrinsic Evaluation Measures for Machine Translation and/or Summarization, pages 65–72.

Shruthi Bannur, Stephanie Hyland, Qianchu Liu, Fernando Perez-Garcia, Maximilian Ilse, Daniel C Castro, Benedikt Boecking, Harshita Sharma, Kenza Bouzid, Anja Thieme, et al. 2023. Learning to exploit temporal structure for biomedical vision-language processing. In Proceedings of the IEEE/CVF Conference on Computer Vision and Pattern Recognition, pages 15016–15027.

James Bergstra, Rémi Bardenet, Yoshua Bengio, and Balázs Kégl. 2011. Algorithms for hyper-parameter optimization. Advances in Neural Information Processing Systems, 24.

Olivier Bodenreider. 2004. The unified medical language system (umls): integrating biomedical terminology. Nucleic Acids Research, 2(uppl_1):D267– D270.

Benedikt Boecking, Naoto Usuyama, Shruthi Bannur, Daniel C Castro, Anton Schwaighofer, Stephanie Hyland, Maria Wetscherek, Tristan Naumann, Aditya Nori, Javier Alvarez-Valle, et al. 2022. Making the most of text semantics to improve biomedical vision– language processing. In Proceedings of European Conference on Computer Vision (ECCV), pages 1–21. Springer.

Kathi Canese and Sarah Weis. 2013. Pubmed: the bibliographic database. The NCBI Handbook, 2(1).

Souradip Chakraborty, Ekaba Bisong, Shweta Bhatt, Thomas Wagner, Riley Elliott, and Francesco Mosconi. 2020. Biomedbert: A pre-trained biomedical language model for qa and ir. In Proceedings of the 28th international conference on computational linguistics, pages 669–679.

Pierre Chambon, Tessa S Cook, and Curtis P Langlotz. 2023. Improved fine-tuning of in-domain transformer model for inferring covid-19 presence in multi-institutional radiology reports. Journal of Digital Imaging, 36(1):164–177.

Peng Chen, Jian Wang, Hongfei Lin, D. Zhao, and Zhihao Yang. 2023. Few-shot biomedical named entity recognition via knowledge-guided instance generation and prompt contrastive learning. Bioinformatics, 39(8):btad496.

Clinical-AI-Apollo. 2023. Clinical-AI-Apollo Medical-NER. HuggingFace.

Jacob Devlin, Ming-Wei Chang, Kenton Lee, and Kristina Toutanova. 2019. Bert: Pre-training of deep bidirectional transformers for language understanding. In Proceedings of the 2019 Conference of the North American Chapter of the Association for Computational Linguistics: Human Language Technologies, Volume 1 (Long and Short Papers), pages 4171– 4186.

Kevin Donnelly et al. 2006. Snomed-ct: The advanced terminology and coding system for ehealth. Studies in Health Technology and Informatics, 121:279.

Hannah Eyre, Alec B Chapman, Kelly S Peterson, Jianlin Shi, Patrick R Alba, Makoto M Jones, Tamara L Box, Scott L DuVall, and Olga V Patterson. 2021. Launching into clinical space with medspacy: a new clinical text processing toolkit in python. In AMIA Annual Symposium Proceedings, volume 2021, page 438.

Christiane Fellbaum. 2010. Wordnet. In Theory and Applications of Ontology: Computer Applications, pages 231–243.

Shlomit Goldberg-Stein, L Alexandre Frigini, Scott Long, Zeyad Metwalli, Xuan V Nguyen, Mark Parker, and Hani Abujudeh. 2017. Acr radpeer committee white paper with 2016 updates: revised scoring system, new classifications, self-review, and subspecialized reports. Journal of the American College of Radiology, 14(8):1080–1086.

Pengcheng He, Jianfeng Gao, and Weizhu Chen. 2022. Debertav3: Improving deberta using electra-style pre-training with gradient-disentangled embedding sharing. In The Eleventh International Conference on Learning Representations.

Pengcheng He, Xiaodong Liu, Jianfeng Gao, and Weizhu Chen. 2020. Deberta: Decoding-enhanced bert with disentangled attention. In International Conference on Learning Representations.

Saahil Jain, Ashwin Agrawal, Adriel Saporta, Steven Truong, Tan Bui, Pierre Chambon, Yuhao Zhang, Matthew P Lungren, Andrew Y Ng, Curtis Langlotz, et al. Radgraph: Extracting clinical entities and relations from radiology reports. In Thirty-fifth Conference on Neural Information Processing Systems Datasets and Benchmarks Track (Round 1).

Albert Q Jiang, Alexandre Sablayrolles, Antoine Roux, Arthur Mensch, Blanche Savary, Chris Bamford, Devendra Singh Chaplot, Diego de las Casas, Emma Bou Hanna, Florian Bressand, et al. 2024. Mixtral of experts. arXiv preprint arXiv:2401.04088.

Qiao Jin, Won Kim, Qingyu Chen, Donald C Comeau, Lana Yeganova, W John Wilbur, and Zhiyong Lu. 2023. Medcpt: Contrastive pre-trained transformers with large-scale pubmed search logs for zero-shot biomedical information retrieval. Bioinformatics, 39(11):btad651.

Alistair Johnson, Lucas Bulgarelli, Tom Pollard, Steven Horng, Leo Anthony Celi, and Roger Mark. 2020. Mimic-iv. PhysioNet. Available online at: https://physionet.org/content/mimiciv/1.0/ (accessed August 23, 2021), pages 49–55.

Alistair EW Johnson, Tom J Pollard, Seth J Berkowitz, Nathaniel R Greenbaum, Matthew P Lungren, Chihying Deng, Roger G Mark, and Steven Horng. 2019. Mimic-cxr, a de-identified publicly available database of chest radiographs with free-text reports. Scientific Data, 6(1):317.

Alistair EW Johnson, Tom J Pollard, Lu Shen, Li-wei H Lehman, Mengling Feng, Mohammad Ghassemi, Benjamin Moody, Peter Szolovits, Leo Anthony Celi, and Roger G Mark. 2016. Mimic-iii, a freely accessible critical care database. Scientific Data, 3(1):1–9.

Vipina K Keloth, Yan Hu, Qianqian Xie, Xueqing Peng, Yan Wang, Andrew Zheng, Melih Selek, Kalpana Raja, Chih Hsuan Wei, Qiao Jin, et al. 2024. Advancing entity recognition in biomedicine via instruction tuning of large language models. Bioinformatics, 40(4):btae163.

Junnan Li, Dongxu Li, Silvio Savarese, and Steven Hoi. 2023. Blip-2: Bootstrapping language-image pre-training with frozen image encoders and large language models. In International Conference on Machine Learning, pages 19730–19742.

Mingchen Li and Rui Zhang. 2023. How far is language model from 100% few-shot named entity recognition in medical domain. arXiv preprint arXiv:2307.00186.

Chin-Yew Lin. 2004. Rouge: A package for automatic evaluation of summaries. In Text Summarization Branches Out, pages 74–81.

Yang Liu, Dan Iter, Yichong Xu, Shuohang Wang, Ruochen Xu, and Chenguang Zhu. 2023. G-eval: Nlg evaluation using gpt-4 with better human alignment. In Processing of the 2023 Conference on Empirical Methods in Natural Language (EMNLP).

Masoud Monajatipoor, Jiaxin Yang, Joel Stremmel, Melika Emami, Fazlolah Mohaghegh, Mozhdeh Rouhsedaghat, and Kai-Wei Chang. 2024. Llms in biomedicine: A study on clinical named entity recognition. arXiv preprint arXiv:2404.07376.

Michael Moor, Oishi Banerjee, Zahra Shakeri Hossein Abad, Harlan M Krumholz, Jure Leskovec, Eric J Topol, and Pranav Rajpurkar. 2023. Foundation models for generalist medical artificial intelligence. Nature, 616(7956):259–265.

OpenAI. Gpt-4v(ision) system card.

OpenAI. 2023. GPT-4 Technical Report. arXiv preprint arXiv:2303.08774.

Kishore Papineni, Salim Roukos, Todd Ward, and Wei-Jing Zhu. 2002. Bleu: a method for automatic evaluation of machine translation. In Proceedings of the 40th annual meeting of the Association for Computational Linguistics, pages 311–318.

Yifan Peng, Shankai Yan, and Zhiyong Lu. 2019. Transfer learning in biomedical natural language processing: An evaluation of bert and elmo on ten bench-marking datasets. In Proceedings of the 18th BioNLP Workshop and Shared Task, pages 58–65.

Pengcheng Qiu, Chaoyi Wu, Xiaoman Zhang, Weixiong Lin, Haicheng Wang, Ya Zhang, Yanfeng Wang, and Weidi Xie. 2024. Towards building multilingual language model for medicine. arXiv preprint arXiv:2402.13963.

François Remy, Kris Demuynck, and Thomas Demeester. 2024. Biolord-2023: semantic textual repre–sentations fusing large language models and clinical knowledge graph insights. Journal of the American Medical Informatics Association, page ocae029.

Akshay Smit, Saahil Jain, Pranav Rajpurkar, Anuj Pareek, Andrew Y Ng, and Matthew Lungren. 2020. Combining automatic labelers and expert annotations for accurate radiology report labeling using bert. In Proceedings of the 2020 Conference on Empirical Methods in Natural Language Processing (EMNLP), pages 1500–1519.

Luca Soldaini and Nazli Goharian. 2016. Quickumls: a fast, unsupervised approach for medical concept extraction. In MedIR Workshop, Sigir, pages 1–4.

Tao Tu, Shekoofeh Azizi, Danny Driess, Mike Schaekermann, Mohamed Amin, Pi-Chuan Chang, Andrew Carroll, Charles Lau, Ryutaro Tanno, Ira Ktena, et al. 2024. Towards generalist biomedical ai. NEJM AI, 1(3):AIoa2300138.

Zilong Wang, Xufang Luo, Xinyang Jiang, Dongsheng Li, and Lili Qiu. 2024. Llm-radjudge: Achieving radiologist-level evaluation for x-ray report generation. arXiv preprint arXiv:2404.00998.

Jerry Wei, Chengrun Yang, Xinying Song, Yifeng Lu, Nathan Hu, Dustin Tran, Daiyi Peng, Ruibo Liu, D. Huang, Cosmo Du, et al. 2024. Long-form factuality in large language models. arXiv preprint arXiv:2403.18802.

Chaoyi Wu, Jiayu Lei, Qiaoyu Zheng, Weike Zhao, Weixiong Lin, Xiaoman Zhang, Xiao Zhou, Ziheng Zhao, Ya Zhang, Yanfeng Wang, et al. 2023a. Can gpt-4v (ision) serve medical applications? case studies on gpt-4v for multimodal medical diagnosis. arXiv preprint arXiv:2310.09909.

Chaoyi Wu, Weixiong Lin, Xiaoman Zhang, Ya Zhang, Weidi Xie, and Yanfeng Wang. 2024. Pmc-llama: toward building open-source language models for medicine. Journal of the American Medical Informatics Association, page ocae045.

Chaoyi Wu, Xiaoman Zhang, Ya Zhang, Yanfeng Wang, and Weidi Xie. 2023b. Towards generalist foundation model for radiology by leveraging web-scale 2d&3d medical data. arXiv preprint arXiv:2308.02463.

Wen-wai Yim, Yujuan Fu, Asma Ben Abacha, Neal Snider, Thomas Lin, and Meliha Yetisgen. 2023. Aci-bench: a novel ambient clinical intelligence dataset for benchmarking automatic visit note generation. Scientific Data, 10(1):586.

Feiyang Yu, Mark Endo, Rayan Krishnan, Ian Pan, Andy Tsai, Eduardo Pontes Reis, Eduardo Kaiser Ururahy Nunes Fonseca, Henrique Min Ho Lee, Zahra Shakeri Hossein Abad, Andrew Y Ng, et al. 2023a. Evaluating progress in automatic chest x-ray radiology report generation. Patterns, 4(9).

Feiyang Yu, Mark Endo, Rayan Krishnan, Ian Pan, Andy Tsai, Eduardo Pontes Reis, EKU Fonseca, Henrique Lee, Zahra Shakeri, Andrew Ng, et al. 2023b. Radiology report expert evaluation (rexval) dataset.

Tianyi Zhang, Varsha Kishore, Felix Wu, Kilian Q. Weinberger, and Yoav Artzi. 2020. Bertscore: Evaluating text generation with bert. In International Conference on Learning Representations.

Xiaoman Zhang, Chaoyi Wu, Ziheng Zhao, Weixiong Lin, Ya Zhang, Yanfeng Wang, and Weidi Xie. 2023. Pmc-vqa: Visual instruction tuning for medical visual question answering. arXiv preprint arXiv:2305.10415.

Ziheng Zhao, Yao Zhang, Chaoyi Wu, Xiaoman Zhang, Ya Zhang, Yanfeng Wang, and Weidi Xie. 2023. One model to rule them all: Towards universal segmentation for medical images with text prompts. arXiv preprint arXiv:2312.17183.

Lianmin Zheng, Wei-Lin Chiang, Ying Sheng, Siyuan Zhuang, Zhanghao Wu, Yonghao Zhuang, Zi Lin, Zhuohan Li, Dacheng Li, Eric Xing, et al. 2024. Judging llm-as-a-judge with mt-bench and chatbot arena. Advances in Neural Information Processing Systems, 36.

Qiaoyu Zheng, Weike Zhao, Chaoyi Wu, Xiaoman Zhang, Ya Zhang, Yanfeng Wang, and Weidi Xie. 2023. Large-scale long-tailed disease diagnosis on radiology images. arXiv preprint arXiv:2312.16151.

Zexuan Zhong and Danqi Chen. 2021. A frustratingly easy approach for entity and relation extraction. In Proceedings of the 2021 Conference of the North American Chapter of the Association for Computational Linguistics: Human Language Technologies, pages 50–61.

Xiao Zhou, Xiaoman Zhang, Chaoyi Wu, Ya Zhang, Weidi Xie, and Yanfeng Wang. 2024. Knowledge-enhanced visual-language pretraining for computational pathology. arXiv preprint arXiv:2404.09942.

